# Care Delivery Gap framework: a proof-of-concept patient-reported measure of guideline-referenced care-process omissions in sickle cell disease

**DOI:** 10.64898/2026.06.08.26355133

**Authors:** Tarimoboere Agbalalah, Adekunle Rowaiye

## Abstract

**Background:** Sickle cell disease (SCD) is concentrated in sub-Saharan Africa, where delivery of guideline-referenced care remains challenging. Current evaluation approaches rely largely on access indicators and clinical outcomes, which do not directly measure care delivery. We developed the Care Delivery Gap (CDG) framework, a patient-reported approach for identifying care-process omissions, and conducted a proof-of-concept study to assess feasibility and explore variation across income strata.

**Methods:** We conducted a cross-sectional framework-development study involving a proof-of-concept sample of 52 individuals with SCD or caregivers recruited through clinics and moderated SCD communities across Africa, North America, and Europe between June 2025 and March 2026. The CDG framework assessed patient-reported omissions in specialist involvement, follow-up continuity, cardiovascular screening, and biochemical surveillance. Analyses were descriptive.

**Results:** Substantial multi-domain care-process omissions were identified despite high reported healthcare engagement. Across geographic income strata, cardiovascular screening was reported by 4/35 (11%) LMIC versus 16/17 (94%) HIC participants, and regular follow-up within the preceding 12 months by 14/35 (40%) versus 16/17 (94%), respectively. High CDG scores, representing omissions across three or four domains, occurred in 20/35 (57%) LMIC compared with 1/17 (6%) HIC participants. Similar disparities were observed across specialist review and vitamin B12 surveillance domains.

**Conclusion:** A structured patient-reported framework identified multi-domain omissions in guideline-referenced SCD care, including among individuals reporting healthcare access. The divergence between access indicators and reported care delivery suggests that service contact alone may not reflect care quality. The framework provides a feasible foundation for future process-level quality measurement in high-burden settings.

## INTRODUCTION

Sickle cell disease (SCD) places its greatest burden on sub-Saharan Africa, where most affected individuals live (1, 2), and where health systems continue to face challenges delivering consistent guideline-referenced care (3-5). Despite this, evaluation of SCD services still relies predominantly on access-oriented indicators such as specialist contact, insurance coverage, and reported follow-up (6, 7). Although widely used as proxies for care quality, these measures do not directly capture delivery of recommended care processes (8). This limitation extends beyond low-resource settings, and commonly used clinical outcomes such as vaso-occlusive crises (VOC) and stroke are influenced by biological heterogeneity, treatment exposure, and healthcare-seeking patterns (6,9-11). These limitations highlight the need for practical approaches to characterising guideline-referenced care delivery at the patient level.

Few existing approaches have used structured multi-domain assessment to measure guideline-referenced SCD care processes. Although exploratory African studies have examined selected deficiencies in SCD care, patient-level evaluation across multiple care domains remains limited, particularly for micronutrient-related and laboratory surveillance processes (12, 13).

Preventive screening, micronutrient assessment, longitudinal monitoring, patient education, and routine follow-up remain inconsistently implemented across healthcare settings, including well-resourced systems (14, 15). In response, we developed a structured Care Delivery Gap (CDG) framework as a pragmatic, guideline-referenced method for identifying patient-reported absence of selected recommended care processes and for capturing whether those processes were reported as completed.

In this proof-of-concept study, we applied the CDG framework in a small purposive multinational sample with an African-centred analytic focus to examine its operational feasibility using patient-reported data. Specifically, we assessed whether reported care-process omissions co-occur with conventional indicators of healthcare access and how patterns of reported care delivery vary across geographic income strata used as sampling categories. These findings provide an initial demonstration of framework applicability and support further refinement in larger populations.

## METHODS

### Study design and participants

We conducted a cross-sectional framework-development survey using a proof-of-concept sample to assess the feasibility of patient-reported measurement of care-process omissions in SCD. Data were collected between June 18, 2025 and March 27, 2026 using a self-administered electronic questionnaire distributed through clinic-based recruitment and moderated SCD-focused digital communities. 52 participants completed the survey. This hybrid strategy was used to capture heterogeneous care experiences, including participants with varying levels of healthcare engagement. A summary of participant recruitment pathways is shown in Supplementary Figure S1. Participants included 43 individuals with self-reported SCD and 9 caregivers of affected children completing the survey on behalf of children under 18 years recruited from Nigeria, Kenya, Uganda, Canada, USA, and UK. Caregiver-completed questionnaires were analysed together with participant self-reports because all survey items related to observable care processes rather than subjective symptom reporting. Recruitment denominator could not be determined because the survey was distributed through multiple community and clinic channels.

### Sample size justification

Because the primary aim was framework feasibility and operational testing rather than prevalence estimation, a formal power calculation was not performed. Consistent with methodological recommendations for pilot and instrument-development studies (16), a sample exceeding 50 participants was considered sufficient to evaluate framework feasibility, item performance, and preliminary score distributions.

### Ethical Consideration

The study was reviewed by the Institutional Review Committee of Cosmopolitan University (CU/ERC/2025/06-1) and deemed exempt from full ethics review under Category 2 (minimal-risk survey research involving human participants). Participation was voluntary and anonymous.

Electronic informed consent was obtained at survey entry after participants received information on study objectives, confidentiality protections, data use, and their right to withdraw at any time without consequence. Survey completion constituted implied consent. No incentives were provided. All data were collected and stored in de-identified form without personally identifiable information.

### Survey variables

The questionnaire collected data on demographic and disease characteristics, healthcare access indicators, reported delivery of selected care processes, and self-reported clinical history. Variables included hematology contact, continuity of follow-up, preventive screening practices, biochemical surveillance, hydroxyurea exposure, VOC frequency, and stroke history. Clinical variables were included for contextual description and were not used as validation anchors for the CDG framework. Geographic grouping was reported descriptively using country income strata (LMIC: Nigeria, Kenya, Uganda; HIC: United Kingdom, United States, Canada) to characterise sampled care environments, based on World Bank Gross National Income per capita classifications (17). The complete questionnaire is provided in Supplementary Table S1.

### Eligibility criteria

Participants were eligible if they had a self-reported diagnosis of SCD or were caregivers completing the survey on behalf of a child with SCD. Individuals living with SCD were included irrespective of age; however, direct survey respondents were required to be aged 18 years or older, able to complete the questionnaire in English, and provide electronic informed consent. Caregiver-completed questionnaires were permitted for participants younger than 18 years. Responses were excluded if questionnaires were incomplete, duplicated, or submitted by individuals without SCD or without caregiving responsibility for a person with SCD.

### Questionnaire and Framework Development

The questionnaire and CDG framework were developed through an iterative multi-stage process. First, a targeted review of SCD clinical practice guidelines and relevant literature was undertaken to identify care processes that were routinely recommended and suitable for patient-report ascertainment. Candidate domains were prioritised according to clinical relevance, feasibility of self-report measurement, applicability across diverse healthcare settings, and potential importance for longitudinal care delivery. Framework development was informed by a patient-investigator with lived experience of SCD, who contributed to domain selection, questionnaire design, item wording, and assessment of patient relevance. Draft items were designed to capture observable care processes rather than subjective perceptions of care quality. Questions underwent iterative review and refinement to improve clarity, minimise ambiguity, and support consistent interpretation by both adult participants and caregivers completing questionnaires on behalf of children. A schematic overview of questionnaire and framework development is provided in Supplementary Figure S2.

### Care Delivery Gap framework

The CDG framework operationalised patient-reported care-process omissions using four predefined domains. Three domains: specialist involvement, continuity of follow-up, and cardiovascular screening, were derived from established SCD guideline recommendations from the National Heart, Lung, and Blood Institute and the American Society of Hematology (18, 19). A fourth domain, biochemical surveillance, was included as an exploratory micronutrient-monitoring indicator based on biological plausibility and emerging interest in nutritional vulnerabilities in SCD rather than a specific guideline recommendation (20). Detailed mappings between framework domains, questionnaire items, eligibility criteria, and source guideline recommendations are provided in Supplementary Table S2. Each unmet domain contributed one point to a composite score ranging from 0–4. Scores were categorised as low (0–1), moderate (2), and high (3–4) levels of care-process omission. Equal weighting was adopted for initial framework simplicity and should not be interpreted as implying equivalent clinical importance between domains. Weighting schemes were intentionally deferred because empirical evidence regarding the relative contribution of individual domains to overall care quality is currently unavailable and will require evaluation in future validation studies. The complete scoring algorithm and category definitions are presented in Supplementary Table S3.

### CDG domain definitions

**Specialist involvement:** no hematology review within the preceding 12 months

**Continuity of follow-up:** no regular clinical follow-up within the preceding 12 months **Biochemical surveillance:** absence of B12 testing and/or unknown status within the preceding 12 months. B12 monitoring was included as an exploratory micronutrient-surveillance domain because chronic hemolytic stress, sustained erythropoietic demand, and multisystem disease burden in SCD may increase vulnerability to nutritional disturbances (20). In addition, micronutrient monitoring in SCD has traditionally focused on folate and iron-related parameters (21, 22), vitamin B12 remains comparatively underexplored despite plausible biological relevance in chronic haemolytic disease. Accordingly, vitamin B12 surveillance was incorporated as a hypothesis-generating care-process indicator rather than a guideline-mandated quality measure.

**Organ-specific preventive screening:** absence of reported cardiovascular screening. This domain was based on participant report of having undergone any heart disease evaluation or cardiovascular assessment, as captured by the questionnaire item **“***Have you been assessed for any heart disease***?**” The questionnaire did not distinguish between specific investigations (e.g., echocardiography, electrocardiography, transcranial Doppler, or specialist cardiology assessment), and responses therefore reflect participant-reported cardiovascular evaluation in broad terms. Cardiovascular screening and continuity of follow-up were additionally examined descriptively because of their relevance to longitudinal SCD care organisation. Age-specific eligibility criteria used for framework application are summarised in Supplementary Table S4.

### Analysis

Analyses were predefined as descriptive. Each CDG domain was analysed individually as a binary indicator and summarised within the composite score. Results are presented as proportions overall and within geographic income strata (LMIC vs. HIC). No formal hypothesis testing or confounder adjustment was performed because analyses were descriptive. No imputation was performed.

Analyses were restricted to completed questionnaires. The study is reported in accordance with STROBE recommendations for observational studies (23).

### Supplementary materials

The Supplement includes questionnaire items used in the present study, detailed mappings between CDG domains and relevant ASH and NHLBI guideline recommendations, domain definitions and scoring rules, the framework-development process, and supplementary tables and figures supporting interpretation of the CDG framework. These materials are provided to enhance transparency, reproducibility, and future refinement of the framework.

## RESULT

### Cohort characteristics

A total of 52 participants were included in descriptive analyses, comprising 43 adults (83%) and nine paediatric participants (17%). Most participants were female (35/52, 67%) and had HbSS genotype (38/52, 73%). The cohort was relatively healthcare-engaged, with 43/52 (83%) reporting tertiary education and 34/52 (65%) reporting insurance coverage or structured healthcare access. Hematology review within the preceding 12 months was reported by 33/52 (63%) participants, while 28/52 (54%) reported regular clinical follow-up during the same interval. Hydroxyurea exposure was reported by 27/52 (52%). High VOC frequency (≥3 crises/year) was reported by 33/52 (63%), and nine participants (17%) reported prior stroke (Table 1).

**Table 1.** Participant Characteristics, Healthcare Access, and Clinical Outcomes (N = 52)

| Characteristic | Total (N=52) |
| --- | --- |
| **Gender** |  |
| Female | 35 (67%) |
| **Age group** |  |
| <18 years | 9 (17%) |
| ≥18 years | 43 (83%) |
| **Genotype** |  |
| HbSS | 38 (73%) |
| HbSC | 10 (19%) |
| HbSβ<sup>0</sup>-thalassaemia | 4 (8%) |
| **Geographic location** |  |
| Nigeria/Kenya/Uganda | 35 (67%) |
| USA/Canada/UK | 17 (33%) |
| **Education** |  |
| Tertiary | 43 (83%) |
| Hydroxyurea use | 27 (52%) |
| Transfusion history | 41 (79%) |
| Healthcare access (insurance/structured care) | 34 (65%) |
| Recent hematology contact (≤12 months) | 33 (63%) |
| Regular follow-up (≤12 months) | 28 (54%) |
| High VOC frequency (≥3/year) | 33 (63%) |
| Stroke history | 9 (17%) |

Data are presented as n (%). All variables were self-reported by participants or caregivers of children with SCD. Healthcare access was defined as self-reported insurance coverage and/or access to structured healthcare services. VOC = vaso-occlusive crisis.

### Reported care-process indicators across geographic strata

Geographic income strata were used solely as descriptive sampling categories and not as proxies for uniform healthcare-system characteristics. Substantial differences in reported care-process omissions were observed across geographic income strata. Among participants from LMIC settings, 18/35 reported no hematology review within the preceding 12 months compared with none of the 17 HIC participants. Similarly, absence of regular follow-up was reported by 20/35 LMIC versus 1/17 HIC participants, while absence of cardiovascular screening was reported by 31/35 and 1/17 participants, respectively (Table 2).

**Table 2.** Care Delivery Indicators by Geographic Income Strata (N = 52)

| Care indicator | LMIC (n=35) | HIC (n=17) |
| --- | --- | --- |
| No hematology review (≤12 months) | 18 (51.4%; 95% CI: 35.9–66.6) | 0 (0%; 95% CI: 0.0–19.5) |
| No regular follow-up (≤12 months) | 20 (57.1%; 95% CI: 41.0–71.9) | 1 (5.9%; 95% CI: 1.1–27.0) |
| No cardiovascular screening | 31 (88.6%; 95% CI: 73.0–95.8) | 1 (5.9%; 95% CI: 1.1–27.0) |
| No vitamin B12 testing/unknown status | 33 (94.3%; 95% CI: 80.8–98.5) | 1 (5.9%; 95% CI: 1.1–27.0) |

Data are presented as n (%; 95% CI). Confidence intervals are provided to illustrate estimate precision and should not be interpreted as population-level prevalence estimates because participants were recruited through purposive clinic-based and community-based sampling. LMIC = low-income and middle-income countries; HIC = high-income countries.

The biochemical surveillance domain demonstrated the greatest disparity between strata. No prior vitamin B12 testing or uncertainty regarding its status was reported by 33/35 LMIC compared with 1/17 HIC participants. Reported receipt of selected care processes showed corresponding differences across strata. Cardiovascular screening was reported by 4/35 (11%) LMIC compared with 16/17 (94%) HIC participants. Regular clinical follow-up within the preceding 12 months was reported by 14/35 (40%) and 16/17 (94%) participants, respectively (Figure 1). Patterns of specialist review also differed substantially between strata. Hematology review within the preceding 3 months was reported by 9/35 (26%) LMIC and 12/17 (71%) HIC participants. Among LMIC participants, an additional 5/35 (14%) reported review within 3–6 months, 3/35 (9%) within 6–12 months, and 10/35 (29%) more than 12 months previously, while 8/35 (23%) reported never having seen a hematologist.

**Figure 1.**
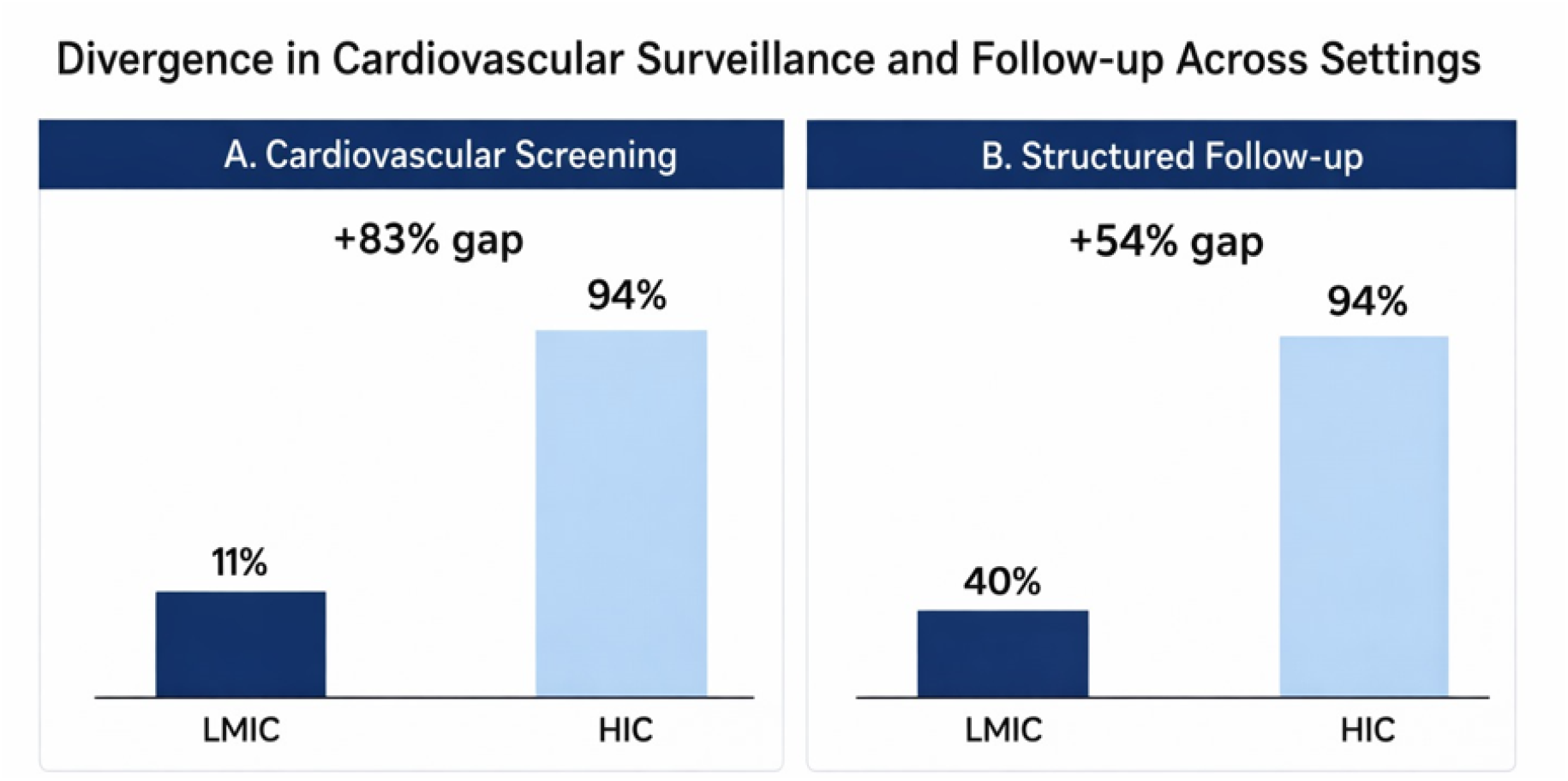
Reported receipt of cardiovascular screening and regular clinical follow-up by geographic income stratum. Bars represent the proportion of participants reporting cardiovascular screening and regular clinical follow-up within the preceding 12 months among participants from LMICs and HICs. Marked differences were observed across sampled care environments, with substantially lower reported receipt of both care processes among LMIC participants. Geographic income strata were used as descriptive sampling categories and should not be interpreted as homogeneous healthcare systems.

### Distribution of CDG scores

High multidomain care-process omission scores (CDG 3–4) were more common among LMIC participants, occurring in 20/35 (57%) individuals, compared with 1/17 (6%) participants from HIC settings. Conversely, low CDG scores (0–1) were observed in 12/17 (71%) HIC participants and 6/35 (17%) LMIC participants. Intermediate scores (CDG = 2) were reported by 9/35 (26%) LMIC participants and 4/17 (24%) HIC participants (Figure 2).

**Figure 2.**
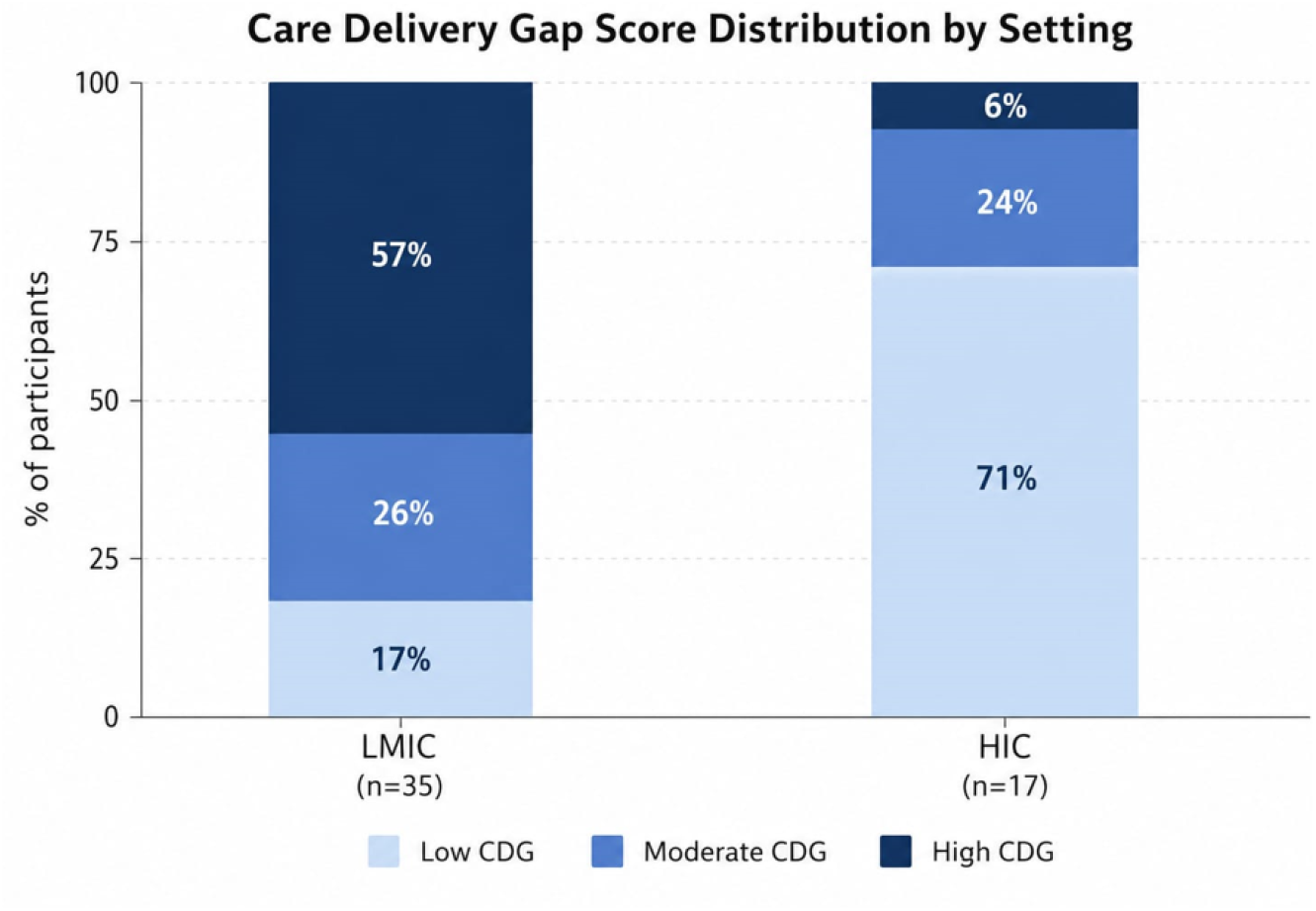
Distribution of Care Delivery Gap scores by geographic income stratum. CDG scores were calculated by summing unmet care-process domains (range 0–4). Scores were grouped as low (0–1), moderate (2), and high (3–4) levels of care-process omission. Higher scores indicate a greater number of reported omissions across assessed domains. Geographic income strata were used as descriptive sampling categories and should not be interpreted as homogeneous healthcare systems.

### Fragmentation and clinical outcomes

CDG scores showed no consistent association with clinical outcomes in this cross-sectional cohort. Participants across the full range of CDG scores reported both low and high frequencies of VOC, with no clear trend observed. Similarly, stroke history did not demonstrate a systematic pattern across levels of care fragmentation. Differences in outcome distribution were observed across study settings. Participants from LMIC exhibited higher levels of reported care fragmentation, while HIC participants reported higher VOC frequency. However, these patterns were not consistent within fragmentation strata and did not indicate a clear relationship between CDG scores and clinical outcomes within or across settings.

## Discussion

This study identified consistent multi-domain omissions in guideline-referenced SCD care processes, including specialist involvement, continuity of follow-up, preventive screening, and biochemical surveillance, even among participants reporting healthcare access. The magnitude of disparities between LMIC and HIC participants was striking: 57% of LMIC participants had high CDG scores (omissions in 3-4 domains) versus only 6% of HIC participants, representing a large effect size (risk difference = 51%). These findings extend prior work documenting SCD care deficiencies in emergency departments and access barriers (24) by demonstrating that care-process gaps persist even among patients with reported healthcare engagement. Our findings align with documented deficiencies in adult SCD care, including poor access to knowledgeable providers and inadequate treatment in emergency settings (24, 25). However, the CDG framework captures routine ambulatory care omissions that conventional service indicators miss. The 94% vs. 11% cardiopulmonary screening disparity between HIC and LMIC participants is consistent with known disparities in SCD care infrastructure documented in prior studies (15, 26).

The biochemical surveillance domain revealed marked variability, with many participants reporting absent vitamin B12 testing or uncertainty regarding B12 status. To our knowledge, micronutrient surveillance has not been routinely incorporated into SCD care-quality frameworks. Because chronic hemolytic stress and sustained erythropoietic demand may increase vulnerability to nutritional disturbances in SCD (27, 28), micronutrient surveillance may represent an important but overlooked care-process domain.

The present findings suggest that the CDG captures dimensions of healthcare delivery that are not readily reflected in commonly reported clinical outcomes such as VOC frequency and stroke history in this cross-sectional cohort. This pattern is not interpreted as a lack of relevance of either construct, but rather as evidence that process indicators and clinical outcomes may represent distinct, partially non-overlapping domains of the SCD care experience (29). This distinction is important because outcomes such as VOC are influenced by multiple factors beyond healthcare delivery, including biological heterogeneity, variation in access to acute care, and differences in outcome reporting (30, 31). Accordingly, the absence of a clear cross-sectional association should not be taken as evidence of independence between constructs, but instead as an indication that CDG and conventional outcomes may capture different aspects of disease burden and healthcare engagement (30). These findings support integrating patient-reported process measures with clinical outcomes in SCD care evaluation (32). However, formal construct validation will require longitudinal studies and linkage with registry and treatment-response data.

### Strengths

This study’s strengths include its multi-continent sampling (Africa, North America, Europe), which is rare in SCD care research; the novel CDG framework providing structured patient-level care-process assessment. Although the observed differences between sampled settings were substantial, the modest sample size and non-probability sampling strategy limit precision and generalisability (19). An additional strength was co-design with a patient-investigator, which supported selection of patient-relevant domains and enhanced interpretability of questionnaire items.

### Limitations

This proof-of-concept study was not designed or powered to estimate population prevalence of care-process omissions. Future multi-site studies using larger and more representative cohorts will be required to evaluate framework performance across diverse SCD populations. Self-report may introduce recall or social desirability bias, though patient-reported data is the only way to capture care-process omissions from the patient perspective. The cross-sectional design precludes causal inference. Recruitment through digital communities may introduce selection bias toward more engaged patients, potentially underestimating true care gaps. The cardiovascular domain used a broad patient-reported measure and did not distinguish between specific screening modalities.

Finally, geographic income strata are descriptive and should not be interpreted as homogeneous healthcare systems.

### Potential implications for future practice and research

Although exploratory, these findings raise broader questions about how SCD care delivery is measured and monitored. The frequent occurrence of reported omissions across multiple domains, including biochemical surveillance and cardiopulmonary screening, suggests that healthcare contact alone may not reliably indicate delivery of recommended care processes. These observations support further evaluation of whether selected care processes are being implemented consistently in routine practice and whether patient-reported process measures can complement conventional access indicators and clinical outcomes in assessing care quality. The substantial differences observed across sampled care environments also highlight the need for deeper investigation of the structural, organisational, and resource-related determinants of care delivery in high-burden settings, particularly across sub-Saharan Africa.

### Future work

Future studies should refine CDG domain definitions, assess reproducibility in larger and more diverse populations, and evaluate integration into routine clinical workflows. Formal psychometric evaluation, including assessment of construct validity, reproducibility, and score behaviour across different clinical contexts, will be required before broader application. Additional work should examine alternative weighting strategies, evaluate inter-domain relationships, and incorporate objective clinical, registry-based, and medical-record data to support external validation and determine whether CDG patterns are associated with disease burden and other clinically meaningful outcomes.

## CONCLUSION

This study demonstrates that patient-reported guideline-referenced care-process omissions can be systematically characterised within a structured multi-domain framework. Further work is needed to evaluate reproducibility, implementation, and utility within clinical audit and quality-improvement contexts. Future integration into SCD programmes and registries may help support process-level monitoring of guideline-referenced care delivery, particularly in African settings.

## Acknowledgements

We gratefully acknowledge all participants who generously contributed their time to complete this questionnaire. We also extend our sincere appreciation to the hematologist who facilitated questionnaire distribution during clinic sessions, thereby enabling participant recruitment.

## Data availability

All data produced in the present manuscript are available upon reasonable request to the authors

## Author Contributions

TA: conceived the study idea, designed the questionnaire, and led manuscript development. AR: Developed figures and visualizations. AJ: Calculated confidence intervals. All authors reviewed the manuscript and approved the final version for submission.

## Generative AI Statement

AI-assisted tools such as Perplexity supported preliminary organization of data elements. However, all data extraction, verification, synthesis, and interpretation were performed manually by T.A. and F.I. to ensure accuracy, methodological rigour, and compliance with PRISMA guidelines. No AI-generated content appears in the final manuscript.

