## Supplemental files for "Care Delivery Gap framework: a proof-of-concept patient-reported measure of guideline-referenced care-process omissions in sickle cell disease"

**Supplementary file**

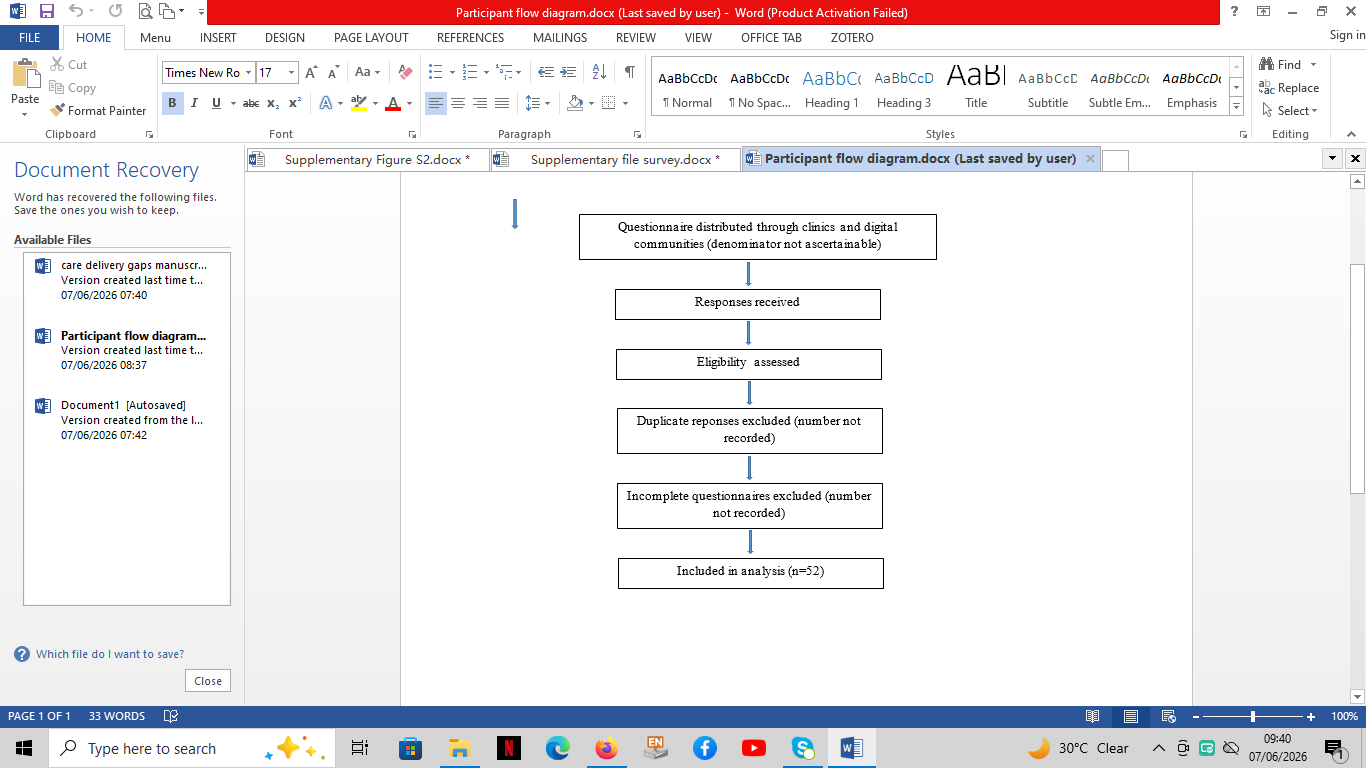

### Supplementary Figure S1: Participant flow diagram

**Supplementary Table S1. Questionnaire items used in the present analysis**

| **Section** | **Questionnaire item** | **Response options** |
| --- | --- | --- |
| Demographics | What is your gender? | Male; Female; Other/Prefer not to say |
| Demographics | What is your age range? | Under 18; 18–25; 26–35; 36–45; 46–55; ≥56 |
| Demographics | What is your country of residence? | Free text |
| Demographics | Education level | No formal education; Primary; Secondary; Tertiary |
| Clinical characteristics | What is your SCD genotype? | HbSS; HbSC; HbSβ-thalassemia; Other |
| Clinical characteristics | When was the last time you had a blood transfusion? | Never; <3 months; 3–6 months; 6–12 months; >12 months |
| Clinical characteristics | How many vaso-occlusive crises have you experienced in the past year? | None; 1–2; 3–5; >5 |
| Clinical characteristics | Have you ever had a stroke or neurological complication? | Yes; No |
| Healthcare access | Do you have health insurance or access to healthcare services? | Yes; No |
| CDG domain | When was the last time you saw a hematologist? | <3 months; 3–6 months; 6–12 months; >12 months; Never |
| CDG domain | Are you currently receiving regular clinic follow-up for SCD? | Yes; No |
| CDG domain | Have you been assessed for heart disease? | Yes; No |
| CDG domain | Do you know your Vitamin B12 levels? | Yes; No; Not sure |

**Abbreviations:** CDG, Care Delivery Gap; SCD, sickle cell disease.

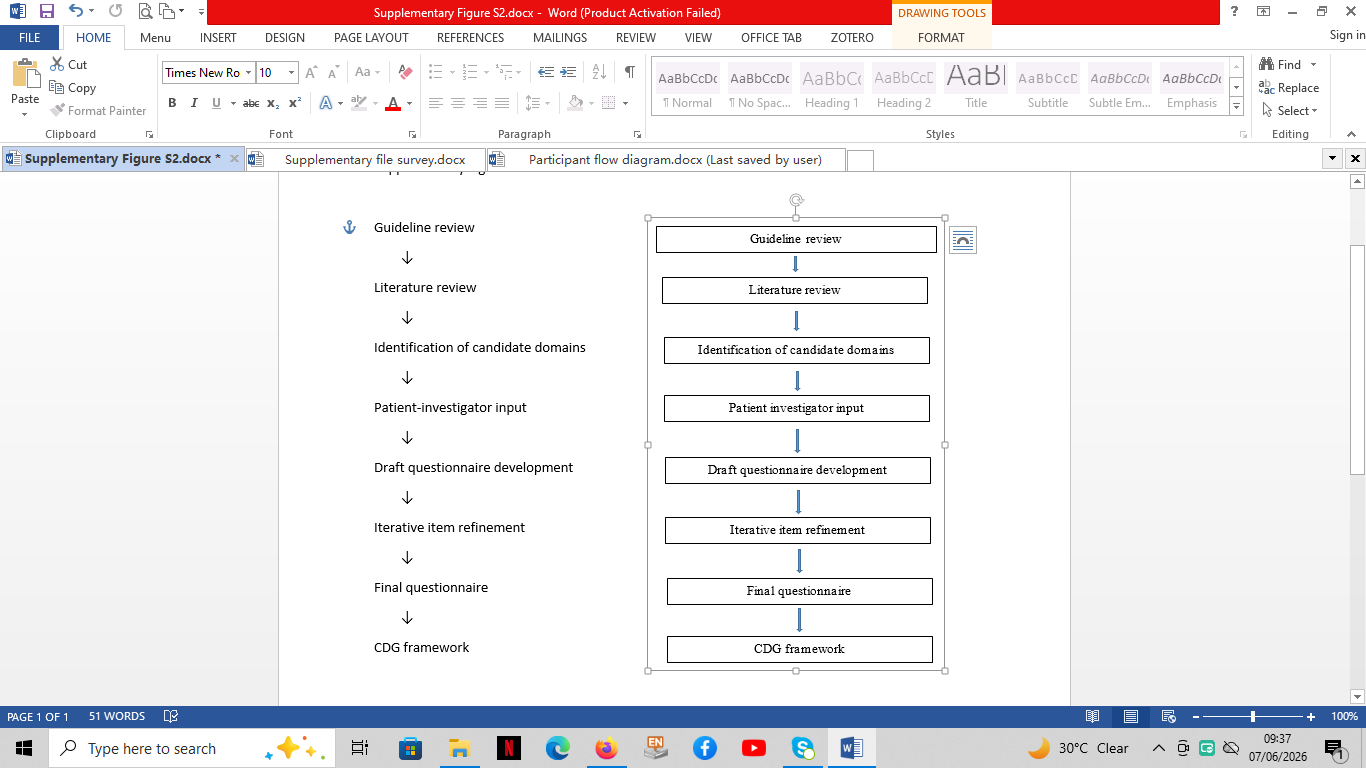

### **Supplementary Figure S2:** Questionnaire and framework development process

**Supplementary Table S2. CDG Framework Domain Mapping**

**Table S2.** Mapping of Care Delivery Gap domains to survey items, scoring rules, and guideline-referenced care processes

| **CDG Domain** | **Survey Question** | **CDG Rule (1 point assigned if unmet)** | **Guideline-Referenced Care Process** | **Primary Guideline Source** |
| --- | --- | --- | --- | --- |
| Specialist involvement | *When was the last time you saw a hematologist?* | No hematology review within the preceding 12 months | Ongoing specialist involvement and access to clinicians experienced in SCD management | NHLBI Expert panel report 2014 |
| Continuity of follow-up | *Are you currently receiving any form of regular clinic follow-up for SCD?* | No regular SCD follow-up within the preceding 12 months | Longitudinal clinical monitoring and routine comprehensive care | NHLBI Expert panel report 2014 |
| Biochemical surveillance* | *Do you know your vitamin B12 levels?* | No reported B12 testing and/or participant uncertain of B12 status within the preceding 12 months | Exploratory micronutrient-surveillance domain included to assess longitudinal monitoring of potentially relevant nutritional parameters in SCD | Exploratory domain; not guideline derived |
| Organ-specific preventive screening | *Have you been assessed for any heart disease?* | No reported cardiopulmonary screening within guideline-referenced intervals | Preventive evaluation for cardiopulmonary complications and organ damage surveillance | ASH 2020 cerebrovascular and cardiopulmonary guidance |

* **Exploratory domain.** Unlike the other three domains, vitamin B12 surveillance was included as a hypothesis-generating micronutrient-monitoring indicator and was not intended to represent a current guideline-mandated quality measure. Its inclusion was based on biological plausibility, chronic hemolytic stress, sustained erythropoietic demand, and the potential overlap between manifestations of vitamin B12 insufficiency and SCD-related clinical features.

**Supplementary Table S3.** Care Delivery Gap scoring algorithm

| **Domain** | **Criterion for domain met (Score = 0)** | **Criterion for domain unmet (Score = 1)** |
| --- | --- | --- |
| Specialist involvement | Hematology review within previous 12 months | No hematology review within previous 12 months |
| Continuity of follow-up | Regular SCD follow-up within previous 12 months | No regular SCD follow-up within previous 12 months |
| Organ-specific preventive screening | Reported cardiovascular/cardiopulmonary screening within guideline-referenced interval | No reported screening within guideline-referenced interval |
| Biochemical surveillance* | Vitamin B12 status known or tested within previous 12 months | No vitamin B12 testing and/or unknown vitamin B12 status |

Exploratory micronutrient-surveillance domain included for hypothesis generation and framework development; not intended as a current guideline-mandated quality indicator.

**Composite score interpretation**

| **Total score** | **Category** |
| --- | --- |
| 0–1 | Low |
| 2 | Moderate |
| 3–4 | High |

**Calculation:** Total CDG score = sum of unmet domains (possible range 0–4). Equal weighting was adopted for proof-of-concept framework development and does not imply equivalent clinical importance across domains.

**Supplementary Table S4:** Age-specific eligibility criteria for CDG domain

| **Domain** | **Adult eligibility** | **Paediatric eligibility** |
| --- | --- | --- |
| Specialist review | All | All |
| Follow-up | All | All |
| Preventive screening | Age-specific if applicable | Age-specific if applicable |
| B12 surveillance | All | All |
